# Topological Data Analysis of Thoracic Radiographic Images shows Improved Radiomics-based Lung Tumor Histology Prediction

**DOI:** 10.1101/2022.05.22.22275410

**Authors:** Robin Vandaele, Pritam Mukherjee, Heather Marie Selby, Rajesh Pravin Shah, Olivier Gevaert

## Abstract

*Topological data analysis* (TDA) provides unparalleled tools to capture local to global structural shape information in data. In particular, its main method under the name of *persistent homology* has found many recent successful applications to both supervised and unsupervised machine learning. Despite its recent gain in popularity, much of its potential for medical image analysis remains undiscovered. In this paper we explore the prominent learning problems on thoracic radiographic images of lung tumors to which persistent homology provides improvements over state-of-the-art radiomic-based learning. It turns out that the novel topological features well capture complementary information important for both ‘*benign* vs. *malignant* ‘ and ‘*adenocarcinoma* vs. *squamous cell carcinoma*’ tumor prediction, while contributing less consistently to ‘*small cell* vs. *non-small cell* ‘—an interesting result in its own right. Furthermore, while radiomic features may be better at predicting malignancy scores assigned by expert radiologists based on visual inspection, it turns out that topological features may be better at predicting the more accurate tumor histology assessed through long-term radiology review, biopsy, surgical resection, progression or response.

## Introduction

The recent rise of quantitative imaging in medicine led to new opportunities for assessing severity, change, and disease through quantifiable features from medical images [1, 2]. In particular, determining lung cancer histology from computed tomography (CT) scan images is a crucial problem in medical image analysis. Machine learning models can lead to rapid diagnosis, intervention, customized treatment, and monitoring of lung cancer patients from such images, reducing the effects of human error in the clinical decision making process. State-of-the-art models are often based on *radiomic features*, which cover a wide range of quantitative tumor characteristics, such as lesion shape, location, and vascularity [3].

Complementary to this, the rising field of *topological data analysis* (TDA) [4], and in particular its main method, *persistent homology* [5], provides an unparalleled tool to quantify local to global structural information in data. Its resulting *persistence diagrams* have been effectively incorporated into learning from topological information for various biomedical machine learning problems. This includes tasks such as predicting protein–protein interaction binding affinity changes [6], biomedical network classification [7, 8], survival prediction of cancer patients [9, 10], and skin lesion segmentation [11].

In this paper we study the use of TDA for lung tumor histology prediction from thoracic radiographic images. Our main goal is to study the added value of TDA to all of the prominent learning problems on lung tumor CT scan images compared to state of the art quantitative imaging tools [12, 13, 14, 15, 16].

This retrospective study was approved by the Institutional Review Board overseeing research at both the VA Palo Alto Health Care System and Stanford University. All CT images were obtained from both the Palo Alto and San Francisco VA picture archiving and communication systems (PACS). We obtained chest CT studies exhibiting cancerous and benign nodules between December 2015 and 2018. For the cancer studies, the inclusion criteria were presence of small cell lung cancer (SCLC), adenocarcinoma (ADC), or squamous cell cancer (SCC). A discussion of the inclusion, exclusion, and size criteria has been previously described [16]. The same criteria were utilized for the San Francisco VA cohort. A CT scan of a primary lung tumor was obtained from each patient, and diagnoses were obtained through biopsy, resection, or serial followup. Table 1 gives an overview of the number of patients per tumor type, with and without added contrast material. Furthermore, the tumor in each scan was manually delineated by an expert radiologist with greater than 10 years of experience using ITK-SNAP [17]. Figure 2a shows an example of the annotation. Figure 1 displays our three classification problems of interest: each pair of siblings with the same parent in the tree induces a binary classification problem.

**Table 1.**
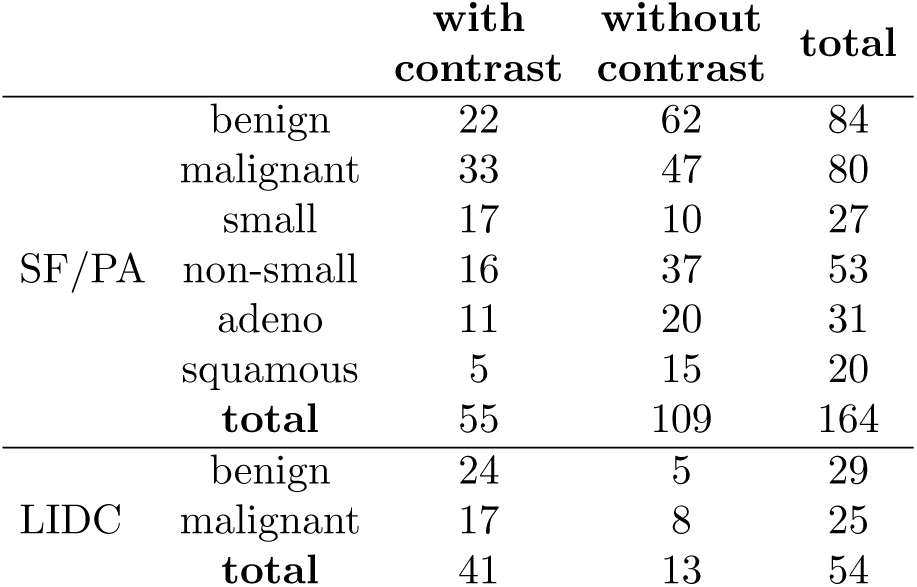
The number of observations for each class of lung tumor in the data, with and without added contrast. Note that the classes in the SF/PA cohort are not mutually distinct (FIGURE 1). Here, the last row thus does not equal the sum of the column values.

**Figure 1.**
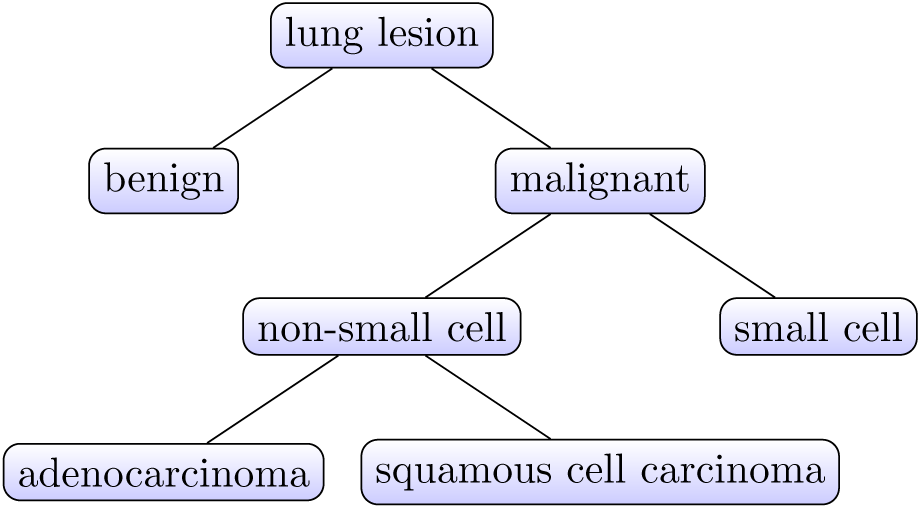
The main hierarchical structure of lung lesions. Each pair of siblings with the same parent in the tree induces a binary classification problem that we study in this paper.

**Figure 2.**
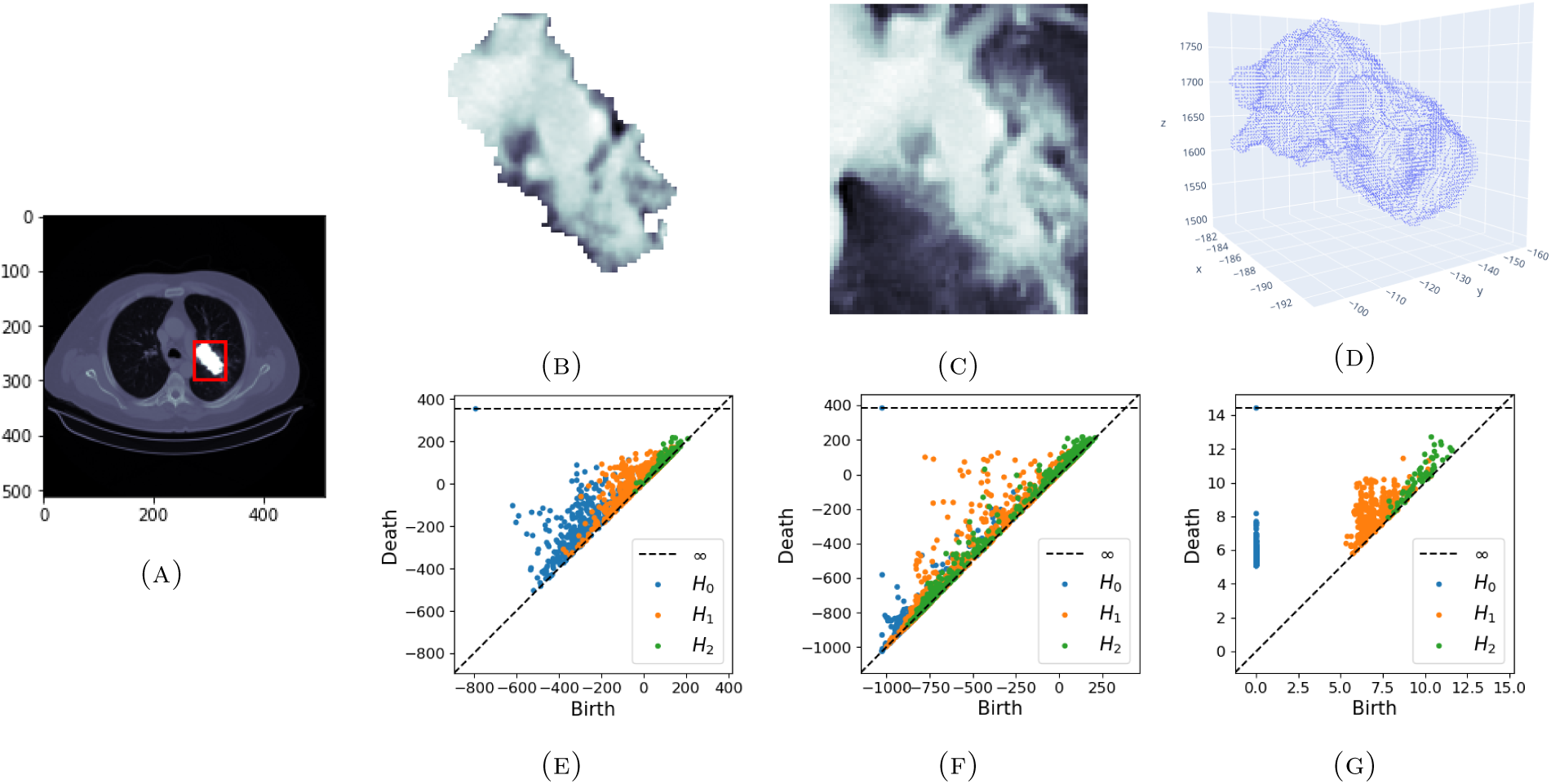
(A) An example slice of a lung tumor CT scan. The tumor in the lungs is marked by a red boundary box. The segmented tumor pixels are in white. (B) A 2D slice of the CT scan image, restricted to the segmented tumor pixels. (C) A 2D slice of the CT scan image, restricted to the pixels in the boundary box of the segmented tumor. (D) A point cloud representing the tumor surface in ℝ^3^. (E) The persistence diagrams obtained from the sublevel filtration of the 3D tumor image. Each group of points with the same color corresponds to one persistence diagram, and captures one dimension of topological hole. (F) The persistence diagrams obtained from the sublevel filtration of the 3D tumor image with surrounding boundary box pixels. (G) The approximated persistence diagrams for the Vietoris-Rips filtration of the point cloud representing the tumor surface in ℝ^3^.

Next, we used CT images from 2544 lung tumor nodules in the Lung Image Database Consortium (LIDC) image collection [18, 19, 20]. These nodules include both primary lung tumors as well as metastatic cancers originating from non-lung tumor sites. The nodules are divided into 807 that were obtained from a scan with contrast material, and 1737 without. The data also contains nodule segmentations provided by multiple expert radiologists for each scan. For tumors with multiple annotations, we used the 50%-consensus segmentation from which we obtained radiomic and topological features, which will be introduced below. Each annotation included a malignancy score on a discrete scale from 1 to 5 assigned by the expert radiologists. The mean of malignancy scores from the different annotations was considered the ‘consensus score’ for each nodule. Besides the malignancy scores, the radiologists also assigned scores for eight semantic features (**sem**): subtlety, internal structure, calcification, sphericity, margin, lobulation, spiculation, and texture.

Finally, we considered the histological diagnosis for benign vs. malignant tumor classification on a smaller LIDC data sample of 54 primary lung tumor nodules, for which true diagnoses were available. These were obtained based on one of the following criteria: review of radiology images to show two years of stable nodule, biopsy, surgical resection, progression or response, [18, 19, 20]. Table 1 summarizes the number of patients per primary tumor type, with and without added contrast material, for which diagnoses were available.

The main contributions of this work are as follows. Through a cohort of lung cancer patients from multiple institutes including Stanford and several VA hospitals, we compare topological features-based classification to standard radiomic features based-classification of lung tumors, with and without contrast material. In particular, we study all of the main lung tumor classification problems that can be considered: ‘benign vs. malignant’, ‘small cell vs. non-small cell’, and ‘adenocarcinoma vs. squamous cell carcinoma’ (Figure 1). We show that topological features consistently provide additional and valuable information when combined with radiomic features for benign vs. malignant classification, and adeno vs. squamous, while interestingly contributing less consistently to small cell vs. non-small cell. Furthermore, the enhanced performance for malignancy prediction is confirmed for both a binary and a continuous outcome on the Lung Image Database Consortium (LIDC) image collection [18, 19, 20]. Finally, we discuss further directions for studying and improving lung tumor histology prediction through TDA. *We emphasize that the technical (mathematical) concepts on which TDA is founded, are omitted from the main paper*. However, a high level and comprehensive introduction to persistent homology and persistence diagrams—from which we derive the topological features—that focuses on medical image analysis of lung tumors, is provided in the supplemental information.

## Results

All results for the various histology prediction problems are summarized in Table 2. Rows correspond to the type of classification or regression problem considered. Columns correspond to the type of features or feature combination that is used: (non-automated) semantic features (**sem**), radiomic features (**rad**), topological features (**top**), concatenated features (**cat**), and a voting (**vote**) or stacking (**stack**) ensemble using both types of features. We also obtained the *p*-value for the null hypothesis that the mean performance when using solely radiomic features is at least as good as using both radiomic and topological features through a voting ensemble. More detailed tables can be found in the supplemental information (Table S1-S10).

**Table 2.** Mean performances in % for lung tumor histology prediction problems with and without contrast (**C**). Best mean performances with automated features are marked in bold.

| problem | C | sem | rad | top | concat | vote | stack | best model | best score | $p$ vote ≥ rad |
| --- | --- | --- | --- | --- | --- | --- | --- | --- | --- | --- |
| benign vs. | Y | - | 84.6 | 86.8 | 86.7 | **87.9** | 88.9 | LR + vote | 88.9 | $5.7 \cdot 10^{-5}$ |
| malignant (SF/PA) | N | - | 74.0 | 75.7 | 76.5 | **78.2** | 73.8 | LR + vote | 80.2 | $1.7 \cdot 10^{-7}$ |
| small cell vs. | Y | - | **77.5** | 62.7 | 66.1 | 75.0 | 71.7 | LR + rad only | 79.8 | 0.94 |
| non-small cell (SF/PA) | N | - | 80.6 | 78.6 | 80.9 | **83.4** | 75.9 | RF + vote | 86.8 | $3.9 \cdot 10^{-2}$ |
| adeno vs. | Y | - | 67.2 | **91.2** | 90.1 | 88.3 | - | RF + top/concat | 98.3 | $1.2 \cdot 10^{-17}$ |
| squamous (SF/PA) | N | - | 64.3 | 70.0 | 68.8 | **71.2** | 65.1 | GNB + vote | 75.0 | $3.8 \cdot 10^{-5}$ |
| malignancy | Y | 61.1 | 56.3 | 52.0 | 53.4 | **59.0** | 53.5 | RF + vote | 61.3 | $5.6 \cdot 10^{-7}$ |
| regression (LIDC) | N | 54.2 | 42.8 | 36.4 | 38.2 | **45.8** | 38.8 | RF + vote | 49.0 | $3.3 \cdot 10^{-9}$ |
| benign vs. | Y | 66.9 | 58.2 | **61.6** | 59.3 | 60.1 | 56.6 | KNN + stack | 67.7 | 0.11 |
| malignant (LIDC) | N | 15.6 | 54.1 | 63.1 | **66.2** | 61.5 | 43.3 | XGB + vote | 78.0 | $1.6 \cdot 10^{-2}$ |

## Lung tumor histology prediction (SF/PA)

We considered three binary classification problems to evaluate TDA and compare it to radiomics-based lung tumor histology prediction: benign vs. malignant, small cell vs. non-small cell, and squamous vs. adenocarcinoma. For benign vs. malignant, we see that adding topological features generally improves solely radiomic features-based classification. In particular, using solely topological features already often outperforms radiomic features for classification. Nevertheless, the voting ensemble using both radiomic and topological features consistently leads to the best performances. Next, for the small cell vs. non-small cell classification problem, we observe that the topological features do not perform as well as radiomic features, both for scans with and without added contrast. However, for images without added contrast, topological features do contribute to the final prediction model through a voting ensemble. Interestingly, both types of features appear to favor scans without contrast material for this particular classification problem. However, the performance differences between the base models with and without contrast are significantly higher for topological features than for radiomic features.

Finally, for the adenocarcinoma vs. squamous cell carcinoma classification problem, we observe the highest performance increases when using topological features, most significantly for images with added contrast. Topological features perform both much better on their own, as well as combined with radiomic features through concatenation or a voting ensemble. Interestingly, there is significantly more performance difference between with and without contrast material for topological features than for radiomic features.

## Lung tumor malignancy prediction from radiologists’ assessment (LIDC)

The outcome here corresponds the continuous malignancy scores assigned by the radiologist for the 2544 lung tumor nodules in the Lung Image Database Consortium (LIDC) image collection, 807 scan with contrast material, and 1737 without. Recall that these were made by the radiologist based on their visual assessments of a set of eight semantic features. For this outcome, the semantic features (which we averaged over different expert annotations) can thus in some sense be considered optimal for prediction. Naturally, there still remains variance in the outcome that is unexplained by solely the semantic features, e.g., due to averaging over the feature and outcome assessments made by different radiologists.

Considering the performances of the automated features, we observe that both with and without contrast material, radiomic features overall perform better than topological features. Thus, the radiomic features appear more applicable to mimic the manual predictions made by the radiologists. Nevertheless, we observe consistent improvements when combining radiomic with topological features through a voting ensemble.

## Lung tumor histology prediction from pathology ground truth (LIDC)

Finally, as discussed above, the manual predictions made by the radiologist are not always representative for the true lung tumor histology. We observe that while radiomic features were better at predicting the radiologist’s manual outcome annotations, topological features are actually better at predicting the accurate tumor diagnoses—at least on this smaller portion of the data for which they are available. Another interesting observation is that while the manually annotated semantic features perform best for images with contrast, they reach poor performance on images without contrast—unlike topological features. A possible explanation is that while the semantic features are valuable for predicting histology, their visual assessment may be more difficult without contrast.

## Discussion

In this paper, we studied the use of topological features for various lung tumor histology classification and regression problems. In particular, we included a thorough overview for all principal prediction problems that might be considered from thoracic radiographic images of the lungs, furthermore splitting our analysis into scans with and without added contrast material.

Our results consistently suggest that TDA provides a promising approach to lung tumor histology prediction from thoracic radiographic images, most notably for ‘benign vs. malignant’ and ‘adeno vs. squamous’ classification. Furthermore, as discussed in the supplemental information, we use a straightforward vectorization through summary statistics to obtain our topological features. Unavoidably, one loses information when transforming topological information into feature representations suitable for machine learning through such (or any other) process. Therefore, a variety of complementary ways to learn through TDA exists. These already found successful machine learning applications in medicine, in particular oncology, for example to predict the survival prognosis of non-small-cell lung cancer patients [9] or to predict the disease-free survival of glioblastoma multiforme brain cancer patients [10]. Thus—even though we already achieved encouraging results in this paper—given the extensive manners to learn from TDA, much of its true potential for rapid diagnosis, intervention, customized treatment, and monitoring of lung cancer patients is yet to be uncovered.

Beyond our main focus on the added effect of TDA for lung tumor histology prediction, our thorough performance evaluations led to extensive quantitative summarizations that are genuine valuable. These include deeply interesting results open to further exploration, such as the different effects when adding contrast material, or the performance differences between semantic, radiomic, and topological features when predicting the malignancy scores assigned by the radiologist vs. when predicting the true tumor histology. For example, we found that while radiomic features may be more suitable for mimicking the radiologists’ malignancy score annotations, topological features may actually be more suitable for predicting the true tumor histology.

## Experimental procedures

### Recourse availability

#### Lead contact

Further information and requests for resources should be directed to the lead contact, Olivier Gevaert.

#### Materials availability

No new materials were generated by this study.

#### Data and code availability

Data to replicate the results summarized in this paper is available fromhttps://github.com/robinvndaele/TDA_LungLesion. This includes persistence diagrams, features, metadata, and outcomes for both the SF/PA and LIDC data. Original scans and masks for the SF/PA cohort are excluded from this repository and are not permitted to be shared. Original LIDC scans and masks are publicly available from https://wiki.cancerimagingarchive.net/display/Public/LIDC-IDRI. All code for this project is available on https://github.com/robinvndaele/TDA_LungLesion.

### Quantitative image features extraction

#### Radiomic features

All images and masks were resampled to 1×1×1 mm3. Radiomic features were then extracted using Pyradiomics 1.0 [21] from the defined regions of interest. We selected 105 Image Biomarker Standardisation Initiative (IBSI)11 compliant features across the following classes: (1) First Order Statistics (19 features), Shape-based (3D) (16 features), Gray Level Size Zone Matrix (GLSZM) (16 features), Gray Level Co-occurrence Matrix (GLCM) (24 features), Gray Level Run Length Matrix (GLRLM) (16 features), Gray Level Size Zone Matrix (GLSZM) (16 features), Neighbouring Gray Tone Difference Matrix (NGTDM) (5 features), and Gray Level Dependence Matrix (14 features).

#### Topological features

From each scan, we obtained different types of *persistence diagrams*. These diagrams, discussed in detail in the supplemental information, quantify topological holes in combinatorial objects termed *simplicial complexes*, which are higher-order generalizations of graphs that are constructed from the scan that grow (thus include more simplices) with some time parameter *t*. This quantification is performed through birth-death pairs (*b, d*), which characterize a topological hole that appeared (*was born*) at time *t* = *b* ∈ ℝ, and that (possibly never) disappeared (*died*), a time *t* = *d* ∈ℝ .∪ {∞}. Topological holes in the lesion, and thus persistence diagrams, can be distinguished by their dimension: *connected components* (dimension 0), *cycles* (dimension 1), and *voids* (dimension 2). These persistence diagrams can furthermore be distinguished by the object through which they capture topological information from the tumor. We considered five such objects, namely the lesion pixels (raw and negated), the lesion pixels with boundary box pixels (raw and negated), and a point cloud representing the lesion surface. Hence, from each scan, we obtained 5 × 3 = 15 persistence diagrams: 3 dimensions of holes for each of the 5 objects. Figure 2 illustrates 3 ×3 examples of such diagrams and the objects they are computed from. Finally, we obtained summarize statistics from these diagrams, as detailed below, resulting in a vector of 290 topological features per scan.

### Supervised machine learning modeling

For each of our machine learning models (discussed hereafter), we used the same feature preprocessing, consisting of the following steps: (1) Missing values (which rarely occurred when a persistence diagram was empty) were imputed with the mean, (2) features were min-max scaled to [0, 1], (3) features were binned into five partitions of equal length (this step serves the following feature selection method which takes discretized features), (4) to maintain a similar number of radiomic and topological features, as well as to reduce the effects of overfitting, we used a feature selection procedure based on minimum redundancy maximum relevance (mRMR) to select 10 features for the final prediction model [22]. We then combined this preprocessing pipeline with each one of six commonly applied classification or regression models (depending on the outcome), namely logistic/linear regression (**LR**), random forest classification/regression (**RF**), *k*-nearest neighbor classification/regression (**KNN**), support vector machine/regressor (**SV**), Gaussian naive Bayes classification/Bayesian regression (**BAY**), and gradient boosted trees classification/regression (**XGB**). Finally, to compare radiomic and topological features, we used various types of feature and model combinations to evaluate the histology prediction models, namely training on radiomic features only (**rad**), training on topological features only (**top**), training on the concatenated features (**concat**), a voting ensemble using models trained on the radiomic and topological features separately (**vote**), and a stacking ensemble trained on the probabilistic output of the models trained on both feature types separately (**stack**). The stacking estimator was equal to the base estimators, e.g., if the base estimators were logistic regression models, so was the final estimator. Note that there were insufficient scans with contrast of squamous tumors in the SF/PA cohort to evaluate a stacking classifier through cross-validation.

### Model evaluation

To evaluate the models and thus the features, we used 10 repeats of 5-fold cross validation to measure the performance of the classification (ROC AUC) and regression (*r*^2^ coefficient of determination) models. Per classification/regression problem, the folds were kept the same over all model evaluations. For the classification problems on the SF/PA data, we used stratified sampling to obtain the folds. This is especially important given the small sample sizes, e.g., to ensure that each fold contains examples of both classes. For the regression problems on the LIDC data, we used standard random sampling. However, this sampling was conducted on the patient level rather than the nodule level, ensuring that different nodules from the same patient were within the same fold. For the classification problems on the LIDC data, we also used stratified sampling. For this, we constructed a class variable indicating whether a patient had a benign tumor nodule (only rarely a patient had both benign and malignant nodules), as to again be able to perform the sampling on the patient level. Note that this class variable was not used for the final outcome, as the lung tumor histology predictions were made on the nodule level.

### Supplemental information description

A high level and comprehensive introduction to persistent homology and persistence diagrams, is provided in the *Supplemental experimental procedures* section. This includes all main concepts required to comprehend the persistence diagrams in Figure 2. This section also discusses the complete topological feature engineering procedures, as well as the hyperparameter settings, that were used to obtain the results discussed in the main paper. Supplementary tables are presented in the *Supplemental items* section.

## Data Availability

Data to replicate the results summarized in this paper is available from https://github.com/robinvndaele/TDA_LungLesion. This includes persistence diagrams, features, metadata, and outcomes for both the SF/PA and LIDC data. Original scans and masks for the SF/PA cohort are excluded from this repository and are not permitted to be shared. Original LIDC scans and masks are
publicly available from https://wiki.cancerimagingarchive.net/display/Public/LIDC-IDRI.

## Acknowledgments

The research leading to these results has received funding from the from the FWO (project no. V407520N, G091017N, G0F9816N, 3G042220), the European Research Council under the European Union’s Seventh Framework Programme (FP7/2007-2013) / ERC Grant Agreement no. 615517, and from the Flemish Government under the “Onderzoeksprogramma Artificiële Intelligentie (AI) Vlaanderen” programme. Next, research reported in this publication was supported by the National Institute of Biomedical Imaging and Bioengineering (NIBIB) of the National Institutes of Health (NIH), R01 EB020527 and R56 EB020527, both to OG. This material is the result of work supported with resources and the use of facilities at the VA Palo Alto Health Care System, Palo Alto, CA. The content is solely the responsibility of the authors and does not necessarily represent the official views of the NIH.

## Author contributions

RV performed the machine learning experiments, topological data analysis, and wrote the main manuscript. PM and HMS provided radiomic features, were included in the discussion of the results, and edited the manuscript. RPS performed data collection and study design. OG supervised the entire project.

## Devlaration of interests

The authors declare no competing interests.

## SUPPLEMENTAL INFORMATION

## Supplemental experimental procedures

### Persistent homology

*Persistent homology* is unarguably the most studied and applied method in topological data analysis (TDA). Its roots are in the field of *algebraic topology* [23], where it has been developed to quantify changes in topological *holes* across a *filtration*, i.e., an ordered sequence of *simplicial complexes*

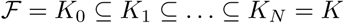

of an initial complex *K*. A simplicial complex *K* can be seen as a generalization of a graph, that apart from nodes (0-simplices) and edges (1-simplices), also includes *higher-dimensional simplices* such as triangles (2-simplices), tetrahedra (3-simplices), …, with the constraint that if *K* contains a simplex *σ*, every simplex *σ′* ⊆ *σ* must also be contained in *K*. Figure S1a illustrates an example of such a filtration.

The topological holes that are quantified through persistence homology, are characterized by their dimension as follows.

- 0-dimensional holes correspond to gaps between connected components.
- 1-dimensional holes correspond to loops, such as the inside of a ring or the handle of a coffee mug.
- 2-dimensional holes correspond to voids, such as the inside of a balloon. In general, a *k*-dimensional hole corresponds to the inside of a *k*-sphere. They can only occur in a space of at least dimension *k* + 1. For *k*≥ 3, these holes become difficult to visualize. These holes are also not used in this paper, since all the mathematical objects (images and point clouds) from which we compute persistent homology occur within a 3-dimensional space.

The number of *k*-dimensional holes of a simplicial complex is denoted by the *Betti-number β*_*k*_. In particular, *β*_0_ denotes the number of connected components.

Given a simplicial complex *K*, and a real-valued function *f* defined on all simplices in *K*, a filtration can generally be written in the form of a *sublevel filtration*

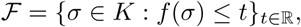

parameterized by a time parameter *t*. In practice, i.e., when dealing with finite data, simplicial complexes in ℱ change for only finitely many values *t*_0_, …, *t* _*N*_ ∈ ℝ. E.g., the filtration in Figure S1a equals the *Vietoris-Rips* filtration, where *K* contains all subsets of a given metric space, i.e., a point cloud dataset, and *f* maps each subset to its diameter. In this case the dataset is 3-dimensional, and hence, no holes of dimension 3 or higher can occur. Thus, we do not include simplices that are higher-dimensional, i.e., include more points, than triangles. The Vietoris-Rips filtration is used to obtain topological information from the point cloud data that models the surface of the lung tumors (Figures 2d and 2g).

**Figure S1.**
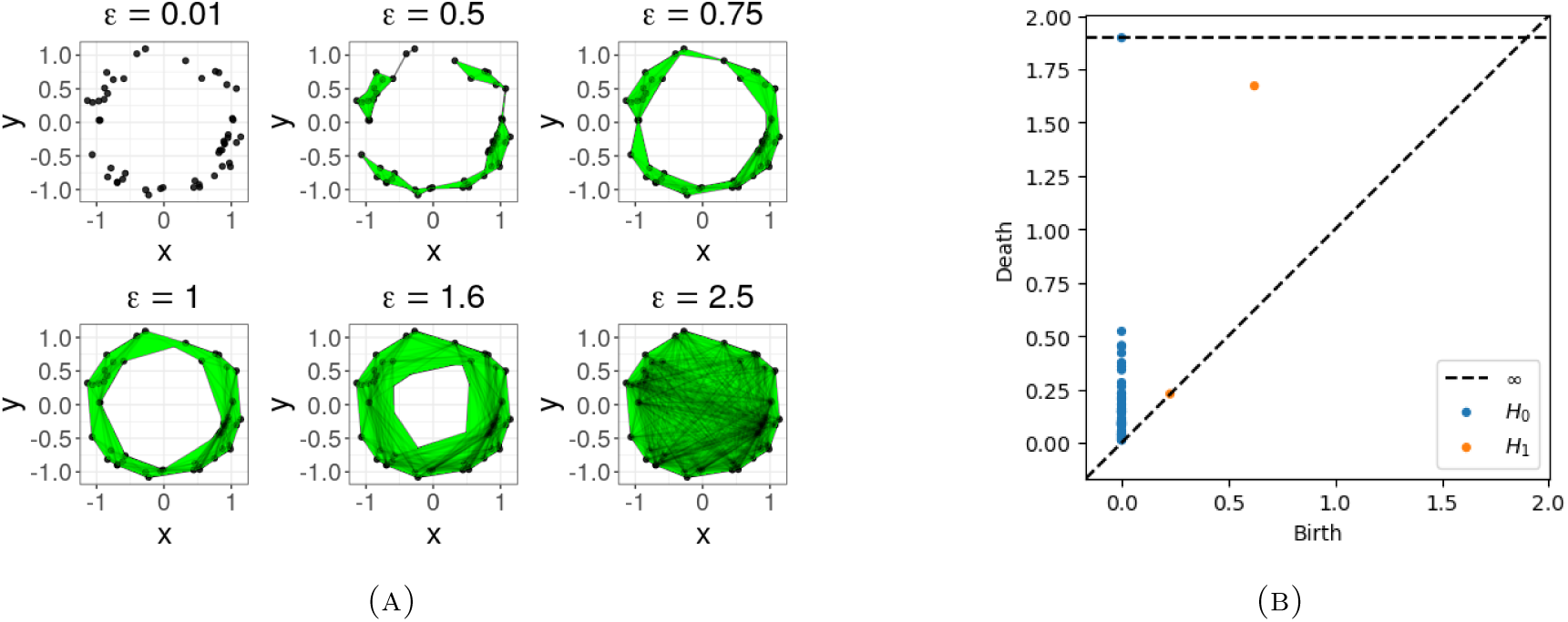
(A) An example of various simplicial complexes in a filtration constructed from a point cloud dataset. Here, the filtration equals the Vietoris-Rips filtration, parameterized by a time (distance) parameter *ϵ*. At time *ϵ*, all simplices with at most three nodes and a diameter of at most *ϵ* are included in the complex. (B) The two corresponding persistence diagrams, one for each considered dimension of hole, plotted on top of each other.

A filtration can also be obtained directly from the CT scan image pixels of a tumor. Note that such scan can be regarded as a 3D array of pixels, of which an example slice restricted to the tumor pixels is shown in Figure 2b. Now any such image naturally defines a simplicial complex *K*, by letting all pixels correspond to 0-simplices. 1-simplices (edges) are then defined by neighboring pixels, and consecutively 2-simplices through the such formed triangles, and 3-simplices through the such formed tetrahedra. The original image can furthermore be regarded as a real-valued function *f* defined on the 0-simplices in *K*, where for a 0-simplex (pixel) *p, f* (*p*) denotes the intensity of *p* in the original image. Through this function, we can define a sublevel filtration directly from the pixel values of the original image as

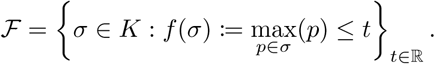

Intuitively, the complex at time *t* is induced by all pixels with intensity at most *t*, and their neighboring relations. The resulting filtration for the image in Figure 2b is shown in Figure S2a. By being inherently 3-dimensional, only up to 2-dimensional holes (voids) can occur in the filtration.

Persistent homology tracks the *birth* (*b*) and *death* (*d*) of these holes across the filtration. The obtained tuples (*b, d*) are then commonly visualized by means of a *persistence diagram*, one for each dimension of hole (Figure S1b). E.g., in Figure S1a, every point defines the birth of a connected component at time *ϵ* = 0. These correspond to the blue points in Figure S1b (H0). By connecting more and more distant points by edges, and ‘filling in’ the resulting triangles, we see that many connected components die (they merge with others). Eventually a 1-dimensional hole (a circle) is formed by the complex, which persists for a relatively long time in the filtration, and finally gets filled in and thus dies. This circle is marked by the highly elevated orange point (H1) in Figure S1b. For the filtration in Figure S2a (of which the persistence diagrams are shown in Figure 2e), the darkest pixels correspond to connected components that are born first. When brighter pixels are consecutively added during the filtration, they may either give rise to new connected components (which are then born as well), or immediately connect to darker pixels which were already present. In the latter case, they may also merge two previously disconnected components, resulting in the death of a connected component. Although 1-and 2-dimensional holes may also be born and die (or even persist indefinitely) during this filtration, this is less apparent from Figure S2a. This becomes more intuitive from Figure S2b.

**Figure S2.**
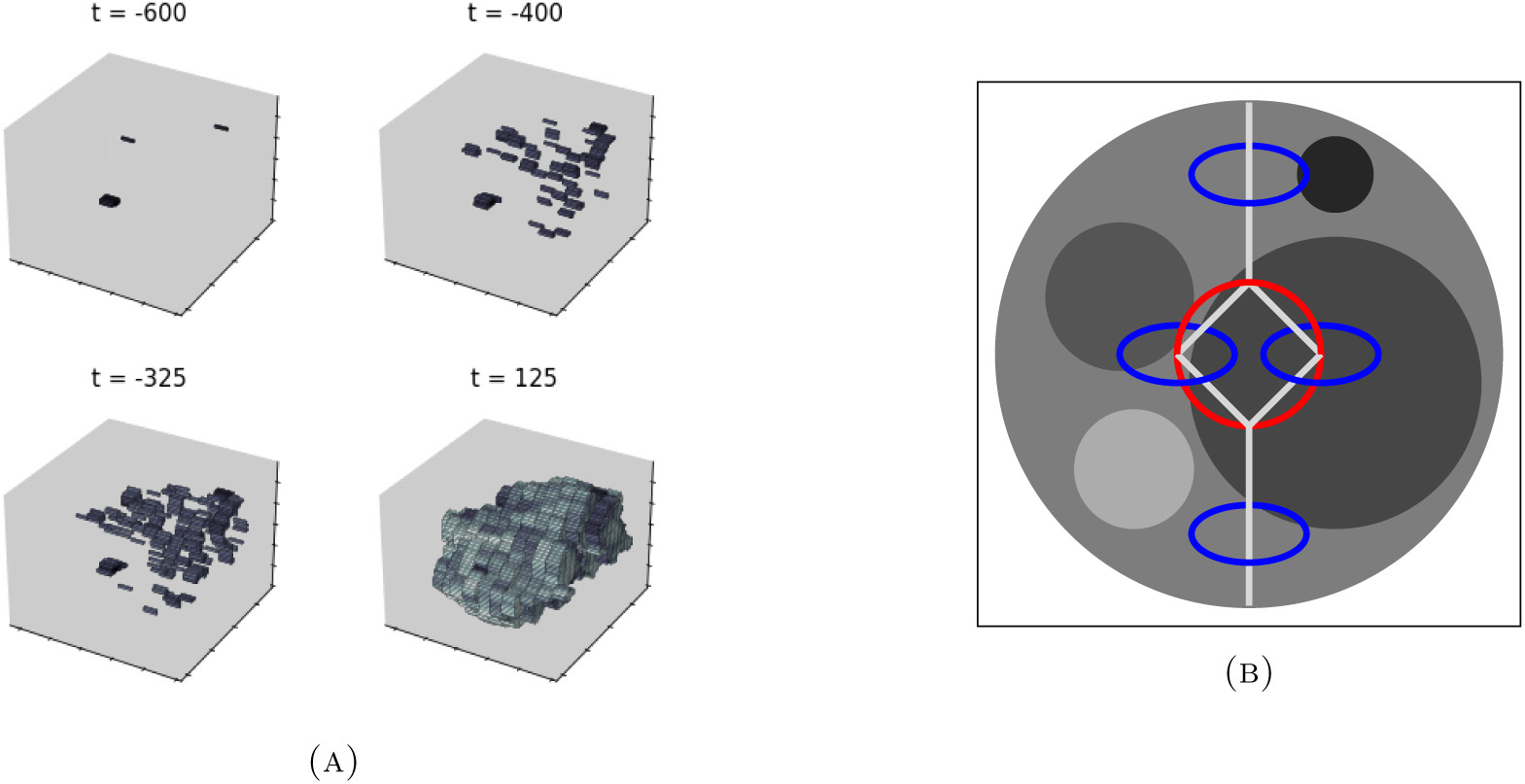
(A) An example of various simplicial complexes in a filtration constructed from the segmented 3D tumor image of which one 2D slice is shown in FIGURE 2a. Here, the filtration equals the sublevel filtration defined by the function *f* mapping each pixel (0-simplex) to its value in the original CT scan image. Darkest pixels are included first. By increasing the time parameter *t*, increasingly brighter pixels are included in the complex as well. 1-, 2-, and 3-simplices are induced by the neighboring relationships between pixels, and not shown in this illustration. (B) An illustration of how persistent homology is able to quantify various topological features directly from image pixels. The illustration should be imagined as representing the restrictions of the holes to a 2D-slice of a 3D-image. Depending on the type of filtration, various structural properties can be inferred trough 1-dimensional persistence, such as holes surrounding vessels when including darker pixels first (blue circles), and holes defined by the vessels themselves when including brighter pixels first (red circle). Hence, different topological information can be obtained from the original and negated pixel values. Various other textural properties, illustrated by the smaller spheres, can also be obtained through 0-dimensional and 2-dimensional persistence.

### Learning from persistence diagrams

One of the main original ideas behind persistent homology and persistence is that holes persisting for a long time–which correspond to ‘highly elevated’ points in their respective persistence diagram—-represent significant features of the underlying topology (hence the name ‘persistence’). E.g., this in Figure S2a the single highly elevated point for connected components (H0) represents that the underlying topological model is connected, and the single highly elevated point for loops (H1) that the underlying topological model contains a cycle.

More recently it has been shown that the entire distribution of points on a persistence diagram may play a significant role in characterizing the data [24, 9, 6]. This is exactly the power of persistent homology for machine learning: it quantifies all of the finest up to the coarsest of topological information in data.

As multisets, persistence diagrams cannot be straightforwardly incorporated into many machine learning models. Various methods have been developed to overcome this issue, as we summarize below.

- Vectorized features of a fixed size can be computed from the persistence diagrams. These can be summary statistics, such as the number of points, or various moments (raw, central, standardized) obtained from their lifetimes. This is the approach we used in this paper. Other examples include Betti curves [25], or binarizations of the persistence diagrams such as persistent images [26].
- Various kernel methods have been developed for learning from persistence diagrams, such as the persistence Fisher kernel [27] and the persistence weighted Gaussian kernel [28].
- Deep learning variants are recently getting more attention [29, 30, 7, 31]. These are designed to learn a task optimal representation of the persistence diagrams at hand.

For a good overview of more general vectorized and kernelized methods that are designed to learn from persistence diagrams, compatible with the scikit-learn library in Python, we recommend [32].

### Filtrations summarizing topological information in lung lesions

From each scan, we obtained the following five to compute topological information from through persistent homology.

1. The sublevel filtration obtained from the raw pixel values, restricted to the segmented lesion (see also Figures 2b and 2e).
2. Same as above, but with negated pixel values.
3. The sublevel filtration obtained from the raw pixel values, restricted to the boundary box of the segmented lesion, which now includes topological information of lung tissue surrounding the lesion (see also Figures 2c and 2f).
4. Same as above, but with negated pixel values.
5. From the given binary lesion segmentation, we first obtained its surface mesh using the marching cubes algorithm [33]. Consecutively, we computed the Vietoris-Rips filtration on the vertices of the mesh, thus a point cloud in the Euclidean space ℝ^3^ (see also Figure 2d). For computational purposes, the persistence diagrams are approximated through the method described in [34] using 1000 landmark points (Figure 2g).

### Persistent homology computation

Computing persistence diagrams was performed in Python using Dionysus (https://pypi.org/project/dionysus/) for image pixels, and Ripser [35] for point clouds.

### Topological features for lung tumor histology prediction

20 summary statistics of the persistence diagrams were collected into a topological feature vector. First, each birth-death pair (*b, d*) was transformed to a *lifespan d b* and a *midlife* 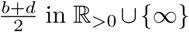 in ℝ_*>*0_− ∪{∞}. These can be interpreted as the prominence, respectively, the location, of a point in the diagram. From each diagram we computed the following statistics.

(1) The minimal birth-time.
(2) The number of infinite lifespans.
(3) The number of finite lifespans.
(4-19) The mean, standard deviation, skewness, kurtosis, first quartile, median, third quartile, and interquartile range of the finite lifespans and finite midlifes.
(20) The entropy of the finite lifespans [36].

Per scan, this thus resulted in a topological feature vector of size 5 × 3 × 20 = 300. However, the number of infinite lifetimes is always the same for the six diagrams obtained from the images with boundary box pixels, as well as for the three diagrams obtained from the point clouds, which all have one 0-dimensional hole and no higher-dimensional holes that persist indefinitely. Similarly, the lowest birth-time in a diagram of the 0-dimensional holes in a point cloud is always 0. These features were thus omitted, resulting in a final vector of 300 − 6 − 3 − 1 = 290 topological features per scan.

We observed that in a few cases there were diagrams without any point (*b, d*) for which *d <* ∞. The summary statistics from the finite lifespans and midlifes are then not straightforwardly defined. In case of the lifespans, we reasoned that any hole that would have been born, died immediately. The statistics for the finite lifespans were then always defined to be 0, analogous to how they would be defined for a random variable that always evaluates to 0. However, an analogous interpretation for the finite midlifes is more difficult. We therefore treated their summary statistics as missing values.

### Hyperparameters and settings

For all considered models, we used their standard settings from the Python libraries scikit-learn and xgboost, apart from changing the output to be probabilistic if needed, i.e., to obtain ROC AUC scores.

### Supplemental items

**Table S1.** ROC AUC performances in % with standard deviations for *benign vs. malignant* classification of lung tumor CT scan images *with added contrast*, using radiomic features (*rad*) and topological features (*top*), as well as for three models combining both: through concatenation (*concat*), soft voting (*vote*), and stacking. Each scores is averaged over 50 models, obtained through 10-repeated stratified samplings in 5 folds. Best scores are marked in bold.

| With contrast |  |  |  |  |  |
| --- | --- | --- | --- | --- | --- |
| model | rad | top | concat | vote | stack |
| LR | 86.7 ± 8.7 | 87.5 ± 10.9 | 87.2 ± 10.7 | **88.9 ± 9.2** | 88.9 ± 9.2 |
| RF | 85.7 ± 12.0 | 87.9 ± 11.0 | 87.5 ± 11.6 | **88.8 ± 11.4** | 86.6 ± 11.6 |
| KNN | 83.9 ± 11.5 | 87.2 ± 10.6 | 87.8 ± 10.0 | **87.9 ± 10.6** | 83.7 ± 10.5 |
| SV | 84.4 ± 9.9 | 85.0 ± 11.6 | 84.4 ± 11.7 | **88.0 ± 9.5** | 87.0 ± 9.6 |
| BAY | 84.9 ± 10.3 | 85.8 ± 11.9 | 86.7 ± 12.1 | **87.6 ± 9.5** | 87.2 ± 9.8 |
| XGB | 81.9 ± 12.8 | **87.6 ± 12.3** | 86.6 ± 13.4 | 86.6 ± 11.6 | 81.5 ± 14.6 |

**Table S2.** ROC AUC performances in % with standard deviations for *benign vs. malignant* classification of lung tumor CT scan images *without added contrast*, using radiomic features (*rad*) and topological features (*top*), as well as for three models combining both: through concatenation (*concat*), soft voting (*vote*), and stacking. Each scores is averaged over 50 models, obtained through 10-repeated stratified samplings in 5 folds. Best scores are marked in bold.

| Without contrast |  |  |  |  |  |
| --- | --- | --- | --- | --- | --- |
| model | rad | top | concat | vote | stack |
| LR | 75.6 ± 10.7 | 77.7 ± 10.0 | 79.0 ± 9.1 | **80.2 ± 9.6** | 79.7 ± 9.5 |
| RF | 72.7 ± 8.8 | 76.3 ± 8.9 | 75.9 ± 9.8 | **77.9 ± 9.3** | 69.5 ± 9.7 |
| KNN | 72.3 ± 10.3 | 75.4 ± 9.8 | 75.5 ± 9.6 | **77.8 ± 9.7** | 72.8 ± 10.1 |
| SV | 74.5 ± 10.8 | 76.2 ± 10.1 | 78.1 ± 9.4 | **78.9 ± 9.9** | 78.5 ± 10.1 |
| BAY | **78.7 ± 9.7** | 74.2 ± 10.6 | 74.7 ± 10.4 | 77.9 ± 9.1 | 76.5 ± 9.8 |
| XGB | 70.0 ± 10.5 | 74.7 ± 11.0 | 75.5 ± 11.0 | **76.5 ± 9.8** | 65.6 ± 12.7 |

**Table S3.** ROC AUC performances in % with standard deviations for *small cell vs. non-small cell* classification of lung tumor CT scan images *with added contrast*, using radiomic features (*rad*) and topological features (*top*), as well as for three models combining both: through concatenation (*concat*), soft voting (*vote*), and stacking. Each scores is averaged over 50 models, obtained through 10-repeated stratified samplings in 5 folds. Best scores are marked in bold.

| With contrast |  |  |  |  |  |
| --- | --- | --- | --- | --- | --- |
| model | rad | top | concat | vote | stack |
| LR | **79.8 ± 18.1** | 62.6 ± 19.7 | 62.1 ± 20.9 | 75.7 ± 19.6 | 74.4 ± 21.1 |
| RF | **78.4 ± 18.8** | 63.2 ± 19.6 | 73.8 ± 17.7 | 76.0 ± 19.4 | 77.2 ± 20.0 |
| KNN | 76.8 ± 17.3 | 64.2 ± 19.5 | 66.8 ± 19.3 | **77.2 ± 18.9** | 74.8 ± 19.1 |
| SV | **79.8 ± 18.1** | 62.6 ± 18.4 | 60.8 ± 20.8 | 75.1 ± 19.5 | 69.1 ± 25.7 |
| BAY | **73.7 ± 20.6** | 62.5 ± 18.4 | 61.5 ± 18.4 | 72.8 ± 19.10 | 64.9 ± 26.5 |
| XGB | **76.4 ± 17.2** | 61.4 ± 22.0 | 71.8 ± 20.6 | 73.3 ± 17.1 | 69.7 ± 22.8 |

**Table S4.**
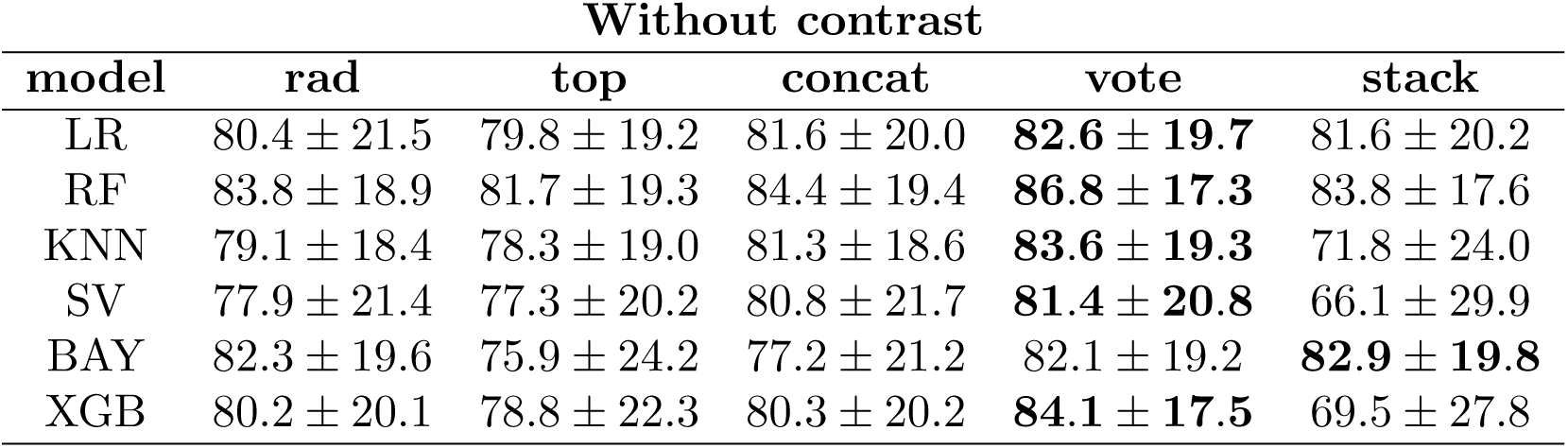
ROC AUC performances in % with standard deviations for *small cell vs. non-small cell* classification of lung tumor CT scan images *without added contrast*, using radiomic features (*rad*) and topological features (*top*), as well as for three models combining both: through concatenation (*concat*), soft voting (*vote*), and stacking. Each scores is averaged over 50 models, obtained through 10-repeated stratified samplings in 5 folds. Best scores are marked in bold.

**Table S5.**
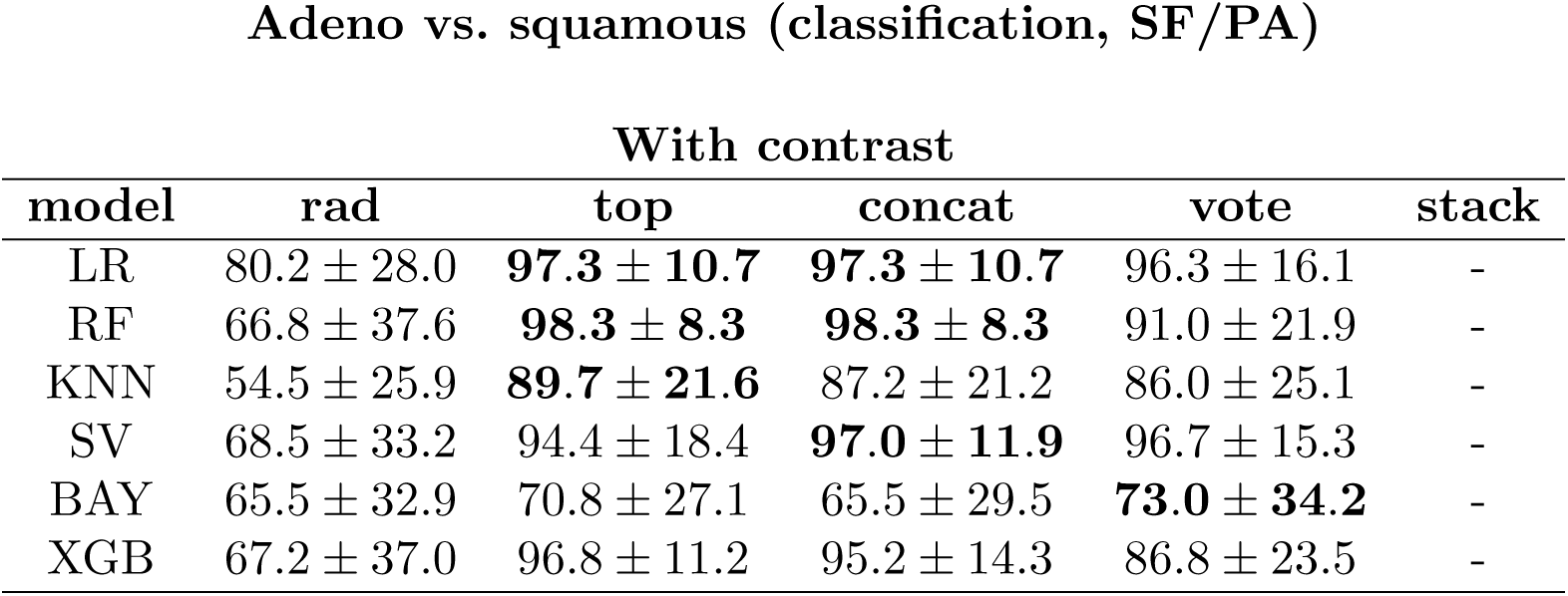
ROC AUC performances in % with standard deviations for *adeno vs. squamous* classification of lung tumor CT scan images *with added contrast*, using radiomic features (*rad*) and topological features (*top*), as well as for three models combining both: through concatenation (*concat*), soft voting (*vote*), and stacking. Each scores is averaged over 50 models, obtained through 10-repeated stratified samplings in 5 folds. Best scores are marked in bold. Note that there were insufficient examples of squamous tumors to train a stacking classifier.

**Table S6.**
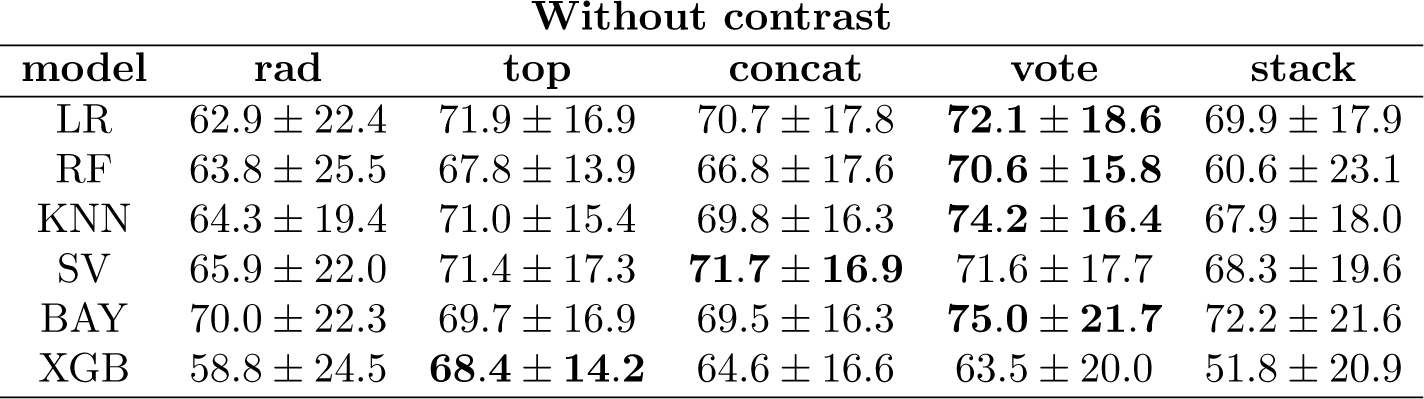
ROC AUC performances in % with standard deviations for *adeno vs. squamous* classification of lung tumor CT scan images *without added contrast*, using radiomic features (*rad*) and topological features (*top*), as well as for three models combining both: through concatenation (*concat*), soft voting (*vote*), and stacking. Each scores is averaged over 50 models, obtained through 10-repeated stratified samplings in 5 folds. Best scores are marked in bold.

**Table S7.** *r*^2^ performances in % with standard deviations for continuous *malignancy* outcome prediction of lung tumor nodules from CT scan images *with added contrast*, using semantic features (*sem*), radiomic features (*rad*) and topological features (*top*), as well as for three models combining both: through concatenation (*concat*), soft voting (*vote*), and stacking. Each scores is averaged over 50 models, obtained through 10-repeated samplings in 5 folds. Non-semantic best scores are marked in bold.

| With contrast |  |  |  |  |  |  |
| --- | --- | --- | --- | --- | --- | --- |
| model | sem | rad | top | concat | vote | stack |
| LR | 61.3 ± 6.4 | 57.5 ± 6.1 | 53.3 ± 5.6 | 53.3 ± 5.5 | 58.5 ± 5.5 | **58.6 ± 5.7** |
| RF | 65.3 ± 6.4 | 57.3 ± 7.0 | 54.1 ± 7.0 | 55.5 ± 6.3 | **61.3 ± 6.1** | 52.6 ± 6.8 |
| KNN | 61.6 ± 6.9 | 57.5 ± 6.4 | 48.7 ± 10.1 | 51.8 ± 8.7 | **59.4 ± 6.5** | 51.9 ± 7.3 |
| SV | 59.9 ± 7.5 | 56.4 ± 7.1 | 51.1 ± 6.0 | 52.4 ± 5.8 | 57.5 ± 6.1 | **57.6 ± 6.5** |
| BAY | 61.4 ± 6.3 | 57.6 ± 5.9 | 53.5 ± 5.6 | 53.6 ± 5.4 | 58.5 ± 5.5 | **58.6 ± 5.8** |
| XGB | 57.0 ± 7.3 | 51.3 ± 9.2 | 51.4 ± 8.0 | 53.5 ± 7.1 | **59.0 ± 7.1** | 41.9 ± 8.4 |

**Table S8.** *r*^2^ performances in % with standard deviations for continuous *malignancy* outcome prediction of lung tumor nodules from CT scan images *without added contrast*, using semantic features (*sem*), radiomic features (*rad*) and topological features (*top*), as well as for three models combining both: through concatenation (*concat*), soft voting (*vote*), and stacking. Each scores is averaged over 50 models, obtained through 10-repeated samplings in 5 folds. Non-semantic best scores are marked in bold.

| Without contrast |  |  |  |  |  |  |
| --- | --- | --- | --- | --- | --- | --- |
| model | sem | rad | top | concat | vote | stack |
| LR | 54.8 ± 4.9 | 43.3 ± 5.5 | 35.6 ± 7.1 | 36.4 ± 6.9 | 44.2 ± 5.2 | **45.1 ± 5.3** |
| RF | 56.9 ± 5.1 | 45.3 ± 6.0 | 41.0 ± 8.0 | 43.6 ± 6.4 | **49.0 ± 5.6** | 36.4 ± 7.9 |
| KNN | 54.2 ± 6.1 | 39.6 ± 7.3 | 31.8 ± 9.9 | 35.4 ± 9.4 | **45.2 ± 6.2** | 34.5 ± 8.2 |
| SV | 54.1 ± 5.1 | 42.1 ± 5.8 | 34.8 ± 7.2 | 35.5 ± 7.3 | 43.8 ± 5.4 | **44.3 ± 5.6** |
| BAY | 54.8 ± 4.9 | 43.5 ± 5.4 | 35.7 ± 7.0 | 36.6 ± 6.9 | 44.1 ± 5.2 | **45.1 ± 5.3** |
| XGB | 50.6 ± 5.3 | 43.0 ± 6.4 | 39.6 ± 8.1 | 41.7 ± 6.6 | **48.3 ± 5.7** | 27.4 ± 11.4 |

**Table S9.** ROC AUC performances in % with standard deviations for *benign vs. malignant* classification of lung tumor nodules from CT scan images *with added contrast*, using semantic features (*sem*), radiomic features (*rad*) and topological features (*top*), as well as for three models combining both: through concatenation (*concat*), soft voting (*vote*), and stacking. Each scores is averaged over 50 models, obtained through 10-repeated samplings in 5 folds. Non-semantic best scores are marked in bold.

| With contrast |  |  |  |  |  |  |
| --- | --- | --- | --- | --- | --- | --- |
| model | sem | rad | top | concat | vote | stack |
| LR | 66.3 ± 20.2 | 53.6 ± 19.3 | **60.3 ± 18.9** | 56.4 ± 17.5 | 57.5 ± 20.2 | 54.2 ± 20.7 |
| RF | 68.2 ± 19.3 | 57.0 ± 18.4 | **61.0 ± 20.2** | 59.3 ± 19.4 | 60.6 ± 18.0 | 50.0 ± 22.1 |
| KNN | 66.8 ± 20.1 | 66.3 ± 17.8 | 61.7 ± 19.0 | 62.4 ± 18.6 | 64.5 ± 18.7 | **67.7 ± 16.4** |
| SV | 67.7 ± 20.8 | 56.7 ± 20.6 | **59.3 ± 19.8** | 57.0 ± 15.0 | 53.9 ± 20.7 | 52.8 ± 19.7 |
| BAY | 64.3 ± 19.5 | 57.4 ± 20.3 | **61.9 ± 19.0** | 59.7 ± 18.8 | 61.3 ± 19.4 | 55.3 ± 22.3 |
| XGB | 68.0 ± 16.3 | 57.9 ± 17.6 | **65.7 ± 17.4** | 61.1 ± 17.8 | 63.0 ± 17.5 | 59.5 ± 21.5 |

**Table S10.**
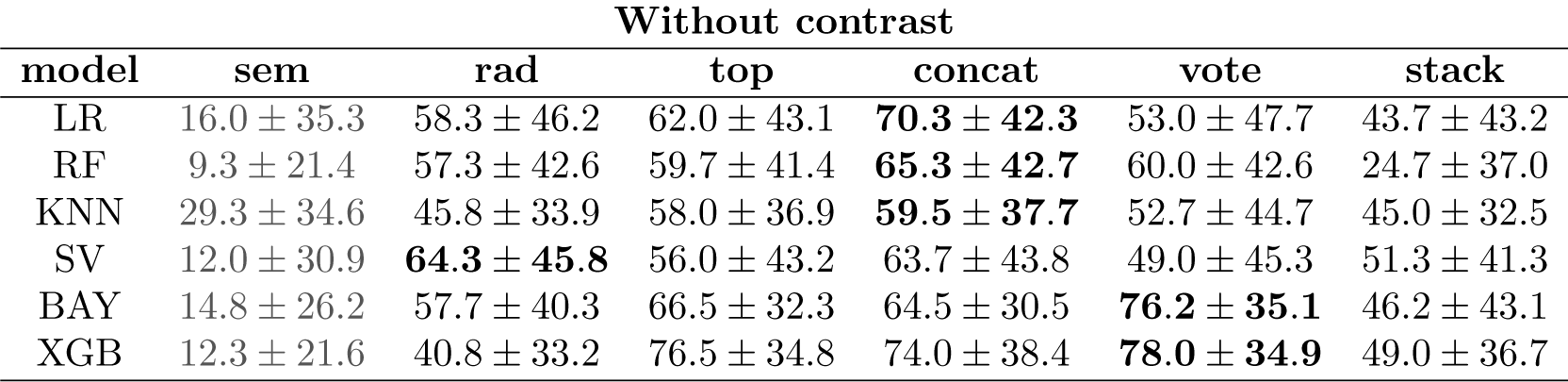
ROC AUC performances in % with standard deviations for *benign vs. malignant* classification of lung tumor nodules from CT scan images *without added contrast*, using semantic features (*sem*), radiomic features (*rad*) and topological features (*top*), as well as for three models combining both: through concatenation (*concat*), soft voting (*vote*), and stacking. Each scores is averaged over 50 models, obtained through 10-repeated samplings in 5 folds. Non-semantic best scores are marked in bold.

